# Diagnostic accuracy of two-photon fluorescence microscopy in the Mohs micrographic surgical margins of squamous cell carcinoma

**DOI:** 10.64898/2026.02.21.26346787

**Authors:** Chi Z. Huang, Vincent D. Ching-Roa, Connor M. Heckman, Katherine Mould, William H. Sipprell, Bruce R. Smoller, Sherrif F. Ibrahim, Michael G. Giacomelli

## Abstract

Cutaneous squamous cell carcinoma (SCC) can be time-consuming to treat with Mohs micrographic surgery (MMS) due to the need for intraoperative frozen section (FS) preparation. Two-photon fluorescence microscopy (TPFM) can generate H&E-equivalent images from fresh tissue specimens in a fraction of this time. To determine the accuracy of TPFM for the evaluation of squamous cell carcinoma in MMS margins compared to conventional FS Mohs slide preparation. TPFM was used to image 144 first stage MMS margins from patients being treated for SCC. A Mohs surgeon reviewed 44 training images and then evaluated 100 margins. After a delay, the same surgeon evaluated the corresponding FS slides. Pairs of TPFM and FS slides were reviewed by an expert dermatopathologist to form a consensus diagnosis. Agreement with consensus diagnosis as assessed by an independent dermatopathologist. 3 margins (3%) unequivocally disagreed with the consensus on TPFM and 2 margins (2%) disagreed on FS. The sensitivity and specificity of TPFM were 95.1% and 98.2%, respectively. This study demonstrates that slide-free histology can be interpreted equivalently to conventional Mohs slide processing by both MMS surgeons and dermatopathologists with minimal training.

## Introduction

Keratinocyte carcinomas (KC) are the most common form of malignancy in the United States, exceeding that of all other cancers combined^1,2^. Of Medicare reimbursed KC treatments, almost half are for squamous cell carcinomas (SCC)^1^. Mohs Micrographic Surgery (MMS) is frequently used to treat cutaneous SCC and achieves the lowest rates of recurrence while conserving healthy tissue through the use of intraoperative complete margin evaluation^3,4^. As a result, MMS obtains better cosmetic outcomes and is preferred for lesions on the face and other anatomic areas where tissue conservation is critical^5^. However, MMS is time-consuming and labor-intensive because histotechnologists must prepare frozen sections (FS) for intraoperative evaluation. This process requires a frozen section lab and takes 20-60 minutes per stage^6^., This can result in long treatment times, inefficient usage of clinic resources, higher treatment costs, and uneven access to offices providing MMS. Furthermore, studies have suggested that artifacts from cryosection can result in a risk of false positives^7,8^, potentially further increasing morbidity and cost.

Reducing both tissue processing times and the labor required for MMS would have a number of advantages. Tissue processing is responsible for a significant fraction of the time patients spend in the clinic. Faster processing would improve clinic resource utilization as well as reduce office time for patients. Furthermore, MMS surgeons frequently operate on multiple patients in parallel due to the delays between stages. Reducing the time for histology processing would ease the complexity of scheduling and treating multiple concurrent patients, as well as the number of procedure rooms and personnel required. Finally, fewer concurrent surgeries may reduce the potential for medical errors by enabling providers to focus on each individual case serially. Slide-free histology methods have emerged as a potentially more efficient alternative to conventional cryosectioning. These methods are faster because they directly image excised tissues without freezing or sectioning, eliminating much of the labor and processing delay while also avoiding freezing artifacts. Furthermore, because no tissue is discarded during the process of sectioning, histology is evaluated at the true surgical margin rather than on a plane below the surface, potentially enabling more accurate evaluation of the true margin status^9,10^. Initial work has demonstrated that confocal microscopy combined with fluorescent stains for DNA can be used analogously to hematoxylin to detect basal cell carcinoma in MMS ^11,12^. Subsequently, dual agent staining using fluorescent analogs of both hematoxylin and eosin was developed^13^. This method can be combined with computational methods to render ‘virtual H&E’ (VH&E) images that closely resemble conventional histopathology, simplifying interpretation and improving sensitivity^14^. This improvement is especially important for SCC which is extremely difficult to interpret without high contrast; consequently, studies using single agent staining for SCC have shown low sensitivity^15^.

A technique related to confocal microscopy, two photon fluorescence microscopy (TPFM), can also utilize multi-agent staining and VH&E rendering to produce images that closely resemble frozen sections^16^. However, unlike confocal microscopy, TPFM uses near-infrared light that penetrates deeply through blood, debris, and tissue, enabling 3D visualization of surgical margins with little preparation. Furthermore, imaging speeds with TPFM are extremely rapid^17^, allowing for fast imaging of even large specimens and multiple specimens in parallel. While our recent study demonstrated promising results using TPFM to detect SCC on simulated biopsies^18^, the accuracy for whole MMS margins has not been assessed. Furthermore, recent advances in fluorescent staining have allowed higher contrast staining, which we hypothesize will further improve sensitivity for SCC^19^. This study investigates a newly developed higher contrast staining method combined with a protocol that enables precise co-registration of whole MMS margins with FS histology^16^. Using these well-aligned TPFM and FS images, we conducted a blinded study analyzing the ability of a MMS surgeon to diagnose SCC on TPFM images as compared to FS on 44 training and evaluation 100 surgical margins.

## Methods

### Specimen collection, preparation, and imaging

Specimens were collected from Rochester Dermatologic Surgery, PC following standard preparation of diagnostic FS sections under a protocol approved by the Research Subjects Review Board at the University of Rochester. In this protocol, following freezing and sectioning to produce diagnostic slides during MMS for SCC, two additional sections were cut from unneeded tissues: a standard 5μm section, which was stained with H&E, and a 50μm thick cryosection used for 3D TPFM imaging. Because both the thick and thin sections are cut back to back, (Figure 1A), they show essentially the same depth into the sample, facilitating co-registration^16^. Samples were stained with 100μg/ml sulforhodamine 101 (SR101, an eosin analog) and 40x (0.4% concentration of stock) SYBR Green (SGr, a hematoxylin analog) in 70% ethanol for 3 minutes, which was sufficient to stain the entire 50 μm thickness. The thin H&E slide was scanned using an OCUS 40 (Grundium Inc.) slide scanner at 40X (0.75 NA) magnification. TPFM imaging was performed using high-speed strip scanning at a 50 MP/s (2.5 mm^2^/s) imaging rate with 250 nm pixels on a microscope system described previously^17^. Briefly, 1040nm excitation light was scanned at 24 KHz line rate using a commercial resonant scanhead (LSK-GR12, Thorlabs Inc.). A 20X/0.7NA air objective with correction collar (UCPLFLN20X, Olympus Corporation) was used to image though the 1mm glass slide to image into the 50μm thick cryosection from the side closest to the matching FS. Detection was performed using high dynamic range silicon photomultipliers (SiPMs), which enable higher imaging rates and sensitivity compared to conventional photomultiplier tubes^20,21^. Two SiPMs were used, one paired with a 525-575nm (Chroma, ET550/50) bandpass for SGr (DNA) and the other a 600-680nm (Chroma, ET640/80) bandpass for SR101 (eosin analog). The high imaging rate enabled an imaging time of 1-3 minutes per level (depending on specimen size), followed by an additional 1-2 minutes for stitching the mosaic images. The TPFM images and scanned slide images were co-registered, and any image pairs with substantial sectioning artifacts in one image but not the other were set aside for the training set. Eleven specimens were excluded due to artifacts or limited co-registration.

**Figure 1:**
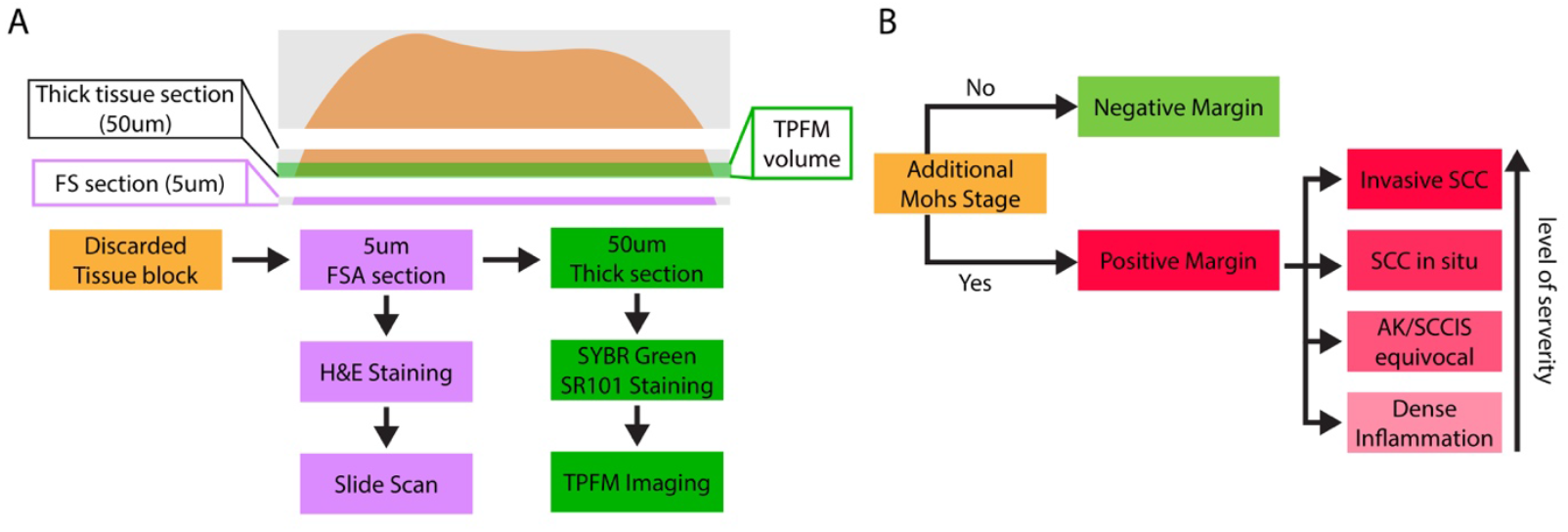
Study workflow and diagnosis options. (A): Sequential FS section and 50μm TPFM imaging volume. The sequential sectioning allows close co-registration between the FS section and the TPFM imaging surface. (B): Treatment based clinically relevant diagnosis options.

### Training and Evaluation by a Mohs surgeon

A practicing MMS surgeon (SFI) reviewed 44 matched pairs of TPFM and H&E frozen section images to gain familiarity with the appearance of SCC under TPFM imaging. Following training, whole slide images and corresponding TPFM images were loaded into a Google Cloud-based comparison program that displayed full resolution, zoomable histology images. The software allowed drawing boxes around areas of pathology and then annotating features observed, including actinic keratosis (AK), SCC and SCC in situ (SCCIS). Additionally, the software allowed recording notes and marking areas as equivocal between actinic keratosis and *in situ* SCC to reflect the subjectivity of classifying the continuous progression between these conditions. Finally, the overall margin status was recorded. We defined a positive surgical margin as any instance where an additional MMS stage would be taken based on the histologic features. Positive histologic features included invasive SCC and SCCIS. The positive margin also included borderline cases between AK and SCCIS or dense inflammation if the reviewer considered the margin equivocal and would take an additional MMS stage (Figure 1B). After a two-week period, corresponding conventional FS images were reviewed in the same software. The time to evaluate each specimen and record the result on TPFM (67s) was not statistically different than FS (62s).

### Consensus Diagnosis

All specimens with a discordant result between TPFM and FS were reviewed by an expert dermatopathologist (BRS) who had not previously seen the images and was not aware of the results of evaluation read. Both TPFM and FS images were reviewed simultaneously, and a consensus diagnosis was developed utilizing the 3D information from the TPFM image stack combined with the FS sections cut adjacent to the TPFM stack. Additionally, any differences due to the few microns difference in level or from sectioning artifacts were recorded.

## Results

### Clinical features

Of the 100 MMS margins in the final evaluation dataset. 42 % of specimens in the evaluation dataset were positive on frozen section (Table 1), with 33 invasive SCC, 8 *in situ* SCC and 3 equivocal between actinic keratosis and SCC *in situ*.

**Table 1.**

| Margin status | Subtype | Number (of 100) |
| --- | --- | --- |
| Positive | Invasive SCC (Well/moderate/poor diff) | 33 |
|  | SCC *in situ* | 8 |
|  | SCCIS/Actinic keratosis equivocal | 3 |
|  | Total | 44 |
| Negative | Total | 56 |

### Agreement between Two Photon Fluorescence Microscopy and FS

Similar histological features were apparent on TPFM VH&E (Figure 2). Accordingly, 92 out of 100 reads agreed exactly. Table 2 summarizes the sensitivity and specificity for detecting SCC by a board-certified fellowship-trained academic MMS surgeon using TPFM. The Kappa value^22^ of 0.839 (CI: [0.732 0.946]) when comparing TPFM to FS suggesting good to very good strength of agreement^23^.

**Table 2.** Sensitivity and specificity of the presence of SCC.

|  |  | False Positive | False Negative |
| --- | --- | --- | --- |
| TPFM vs Consensus |  | 1 | 4 |
| TPFM vs Consensus<br>(Exclude Equivocal) |  | 1 | 2 |
| FS vs Consensus |  | 0 | 2 |
|  | Sensitivity | Specificity | Accuracy |
| TPFM | 90.7%<br>[77.8%, 97.4%] | 98.2%<br>[90.5%, 100%] | 95.0%<br>[88.6% 98.3%] |
| TPFM<br>(Exclude Equivocal) | 95.1%<br>[83.5%, 99.4%] | 98.2%<br>[90.5%, 100%] | 96.9%<br>[91.2% 99.4%] |

**Figure 2:**
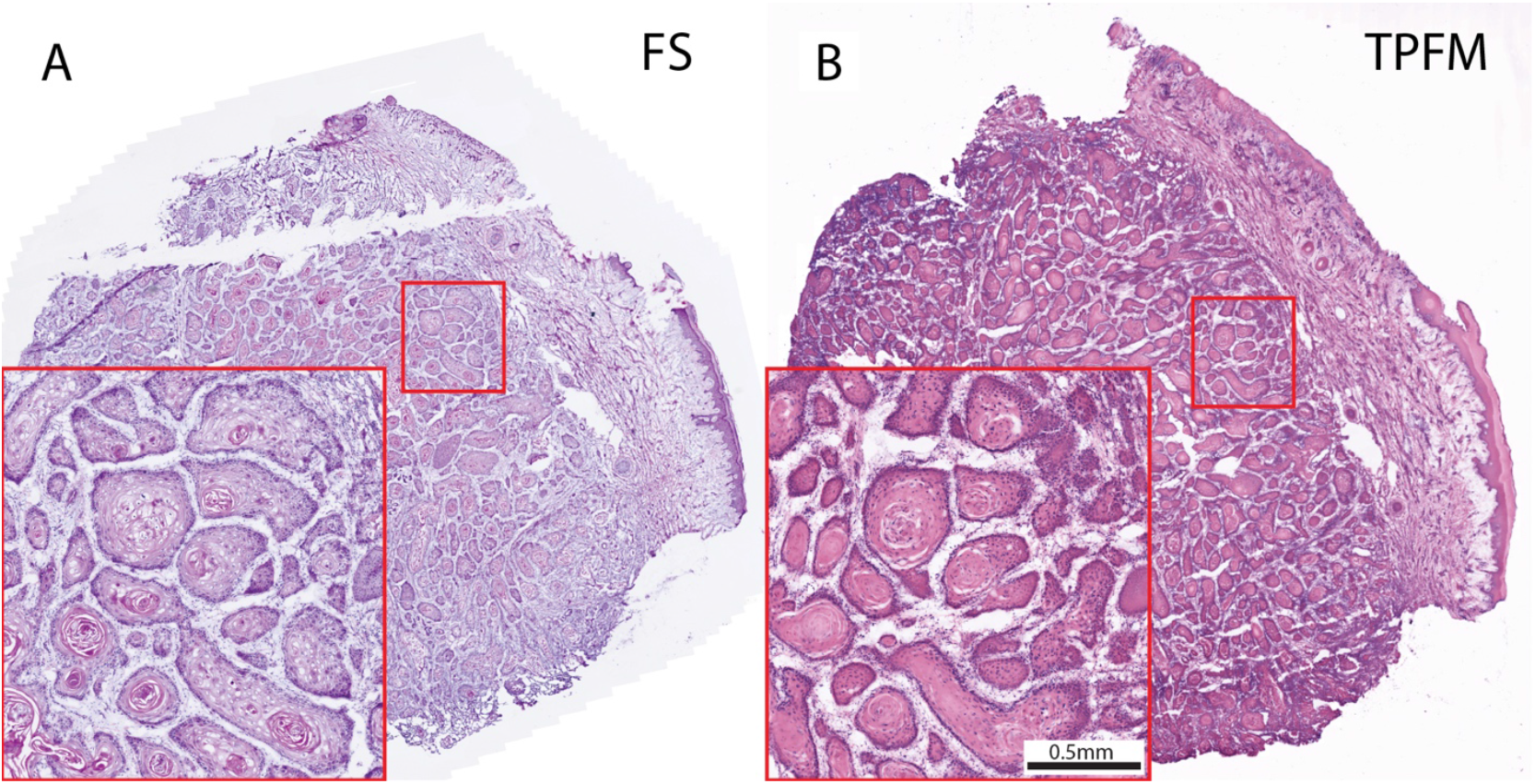
Well-differentiated squamous cell carcinoma (A&B) MMS surgical margin. (A): Full field brightfield frozen section image of a well-differentiated squamous cell carcinoma. (B): The corresponding TPFM image.

### Consensus diagnosis and classification of discordances

The 8 samples that did not agree between TPFM and FS were reviewed to determine a consensus diagnosis and classify the discordance. On two samples, small areas of SCC (one invasive and one *in situ*) were identified on both the TPFM and FS images by the expert reader but were not identified during the evaluation read of the FS (Figure 3A). We classify these two samples as false negative on FS (Table 2). There was a single instance of invasive SCC that was identified by the expert reader but was not identified during the evaluation read on TPFM (Figure 3B). We classify this sample as a false negative on TPFM. One margin contained dense inflammation that was judged to contain SCC on deeper TPFM sections but not to contain SCC on the more superficial FS, which we conservatively classify as a false positive on TPFM. Finally, one sample was actinic keratosis on TPFM but SCC *in situ* on FS, which we classify as a false negative.

**Figure 3:**
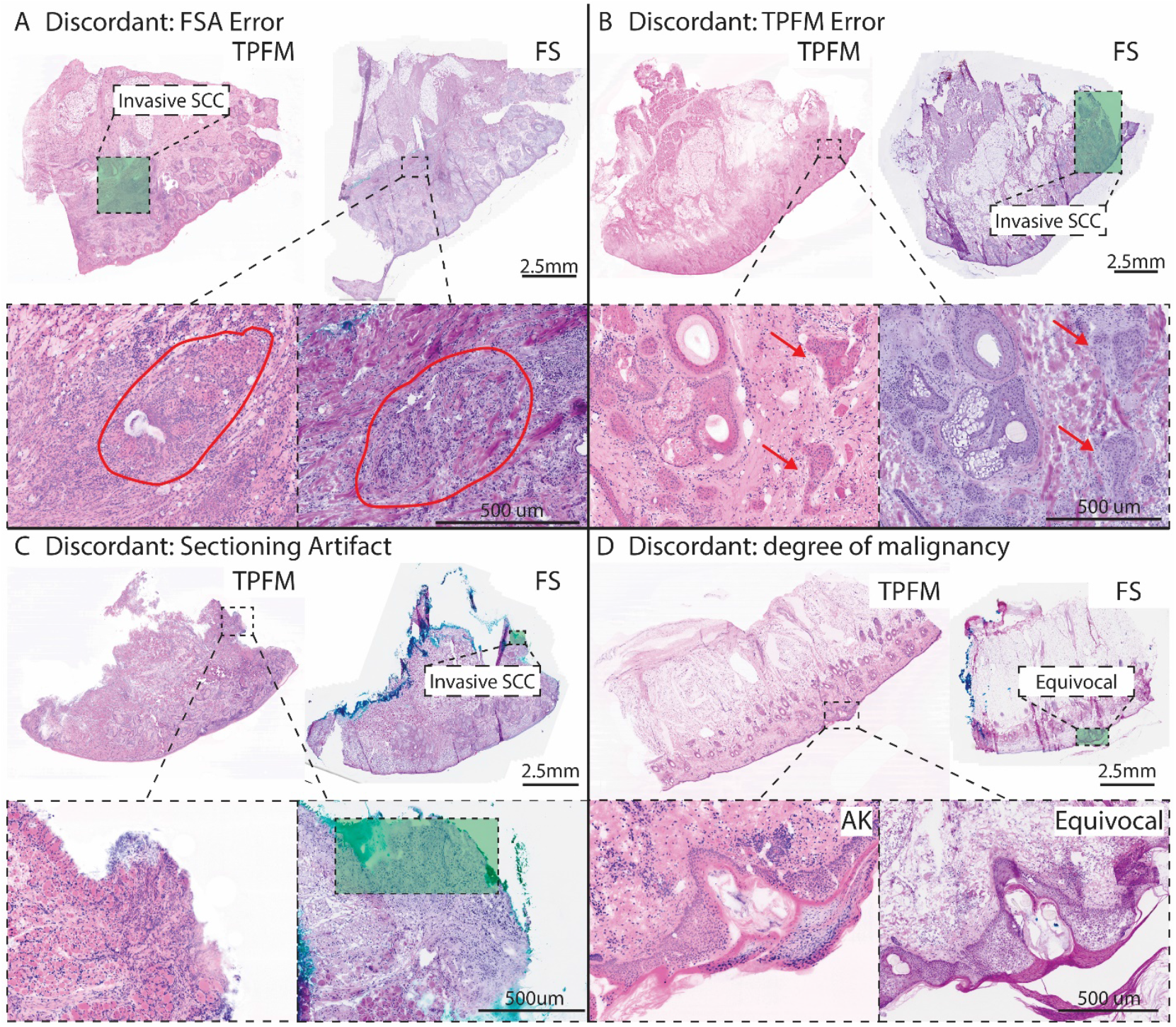
Analysis of discordant samples. The green box indicates locations labeled by the reader as containing pathology. A) missed pathology on FS, B) missed pathology on TPFM, C) tumor lost to sectioning artifact, and D) a sample that disagreed on the degree of malignancy between an actinic keratosis and equivocal between actinic keratosis and SCC in situ on FS.

One sample had a sectioning artifact where a small focus of tumor was present in the FS, but the area was damaged and not evaluable in the TPFM image (Figure 3C). We excluded this sample from analysis because the area marked positive on FS was not interpretable on TPFM due to artifact of cryosectioning.

Finally, in the remaining 2 samples, the disagreement could not be definitively classified as an error in either FS or TPFM due to disagreement on how to classify the continuous progression from actinic keratosis to *in situ* SCC (Figure 3D). In both cases, the expert reader agreed with the equivocal diagnosis but favored the TPFM diagnosis in one and favored the FS diagnosis in the other. As these could not be definitively classified, we analyzed them separately.

### Overall sensitivity and specificity of TPFM for evaluation of SCC

To calculate the overall sensitivity and specificity of TPFM, the single sample where the tumor was obscured by sectioning artifact was excluded from analysis, as none of the small areas annotated as SCC was present on the TPFM image. Furthermore, the two discordances caused by errors on the FS read were counted as true positives for TPFM. The two equivocal samples were conservatively assumed to be false negatives on TPFM as shown in Table 2. The sensitivity was 90.7% (95%CI: 77.8% to 97.4%), and the specificity was 98.2% (95%CI: 90.5% to 100%). Conversely, if the two disagreements that were equivocal and thus could not be definitively concluded to be in disagreement are excluded, sensitivity improves to 95.1% (CI: 83.5% to 99.4%) and specificity remains the same at 98.2% (CI: 90.5% to 100%). In comparison, the FS had close to identical sensitivity to TPFM (95.4% CI: 84.2% to 99.4%) with slightly higher specificity (100% CI: 93.5% to 100%) when compared to the actual read during MMS.

## Discussion

Slide-free histology has a number of advantages over conventional frozen section processing, including faster processing times, freeing urgently needed^24,25^ histotechnologists for other tasks, and the ability to assess the true surgical margin, rather than a deeper surface cut closer to the tumor core by cryosectioning^7,8^. More subtle advantages include the elimination of sectioning artifacts, newer fluorescent stains that have much higher contrast than conventional H&E labeling, as well as the ability to image multiple planes in 3D (“optical sectioning”) that may simplify interpretation and reduce errors due to ambiguous or subtle pathology. Accordingly, in this evaluation of 100 first stage MMS excisions that were precisely co-registered with the FS cutting plane to minimize errors due to co-registration, we observed two instances where pathology was overlooked on FS that were correctly identified on TPFM. If disagreements between samples that were equivocal on FS are excluded, there were 3 discordances on TPFM, although 2 of these represented differences of degree of malignancy present, and only a single false negative was an unambiguous error in which definitive pathology was overlooked. Combined with a recent case series demonstrating TPFM imaging in a Mohs clinic^26^, these results strongly suggest that TPFM could be utilized in place of FS for the evaluation of MMS margins during surgery for SCC.

The overall sensitivity and specificity of TPFM including equivocal samples was 90.7% and the specificity was 98.2%, respectively. Sensitivity improves to 95.1% if disagreements on equivocal samples (where it is unclear if the FS or TPFM diagnosis is correct) are excluded. These results greatly outperform the limited studies of slide-free histology for SCC margins published previously, the largest of which found a sensitivity of only 50% for SCC on MMS margins^15^. A limitation of previous work that may account for the large difference in sensitivity is the use of acridine orange as a single-agent stain to label both stroma and DNA. In contrast to newer cyanine-derived stains such as SYBR Green, the low specificity and high background staining of these agents combined with the lack of an eosin-analog counterstain makes interpretation of dense epithelial tissues extremely challenging. Conversely, the protocol we demonstrate here produces high quality images that closely reproduce the appearance of conventional H&E staining, is inexpensive, does not interfere with subsequent immunohistochemistry, and enables highly accurate interpretation of SCC with minimal training.

Limitations of this study include the involvement of a single site and single Mohs surgeon and postoperative evaluation of cryosectioned tissues rather than fresh surgical margins. Future studies will be needed to evaluate any possible improvements in treatment accuracy or decreases in treatment times associated with slide-free histology in actual surgical practice and to investigate other malignancies such as melanoma or Merkel cell carcinoma. A similar study investigating the use of TPFM in basal cell carcinoma is in progress

## Conclusion

TPFM can evaluate first stage MMS margins during surgery for SCC with equivalent accuracy to FS. Future studies are needed to assess if patient care and treatment times can be improved.

## Data Availability

All data produced in the present study are available upon reasonable request to the authors

## IRB approval status

Reviewed and approved by the University of Rochester Research Subjects Review Board; STUDY00003085

## References

1. Rogers HW, Weinstock MA, Feldman SR, Coldiron BM. Incidence Estimate of Nonmelanoma Skin Cancer (Keratinocyte Carcinomas) in the US Population, 2012. JAMA Dermatology. 2015;151(10):1081–1086. doi:10.1001/jamadermatol.2015.1187

2. Siegel RL, Giaquinto AN, Jemal A. Cancer statistics, 2024. CA: A Cancer Journal for Clinicians. 2024;74(1):12–49. doi:10.3322/caac.21820

3. Leibovitch I, Huilgol SC, Selva D, Richards S, Paver R. Basal cell carcinoma treated with Mohs surgery in Australia II. Outcome at 5-year follow-up. Journal of the American Academy of Dermatology. 2005;53(3):452–457. doi:10.1016/j.jaad.2005.04.087

4. van Loo E, Mosterd K, Krekels GAM, et al. Surgical excision versus Mohs’ micrographic surgery for basal cell carcinoma of the face: A randomised clinical trial with 10 year follow-up. Eur J Cancer. 2014;50(17):3011–3020. doi:10.1016/j.ejca.2014.08.018

5. Celoria V, Rosset F, Pertusi G, et al. Advantages of Mohs Surgery in the Treatment of NMSC in the Head and Neck District. Journal of Clinical Medicine. 2025;14(13):13. doi:10.3390/jcm14134732

6. Gauthier P, Ngo H, Azar K, et al. Mohs surgery - a new approach with a mould and glass discs: review of the literature and comparative study. J Otolaryngol. 2006;35(5):292–304. doi:10.2310/7070.2005.4047

7. Varma S, Boitor R, Notingher I. Achieve Early Complete Sections in Mohs Surgery or Beware of False Positives. Dermatologic Surgery. 2021;47(6):832. doi:10.1097/DSS.0000000000002844

8. Taylor BR, Groover JA, Cook J. Facing the Block and False Positives in Mohs Surgery: A Retrospective Study of 2,198 Cases. Dermatologic Surgery. 2013;39(11):1662. doi:10.1111/dsu.12341

9. Huang CZ, Ching-Roa VD, Heckman CM, et al. Piston-based specimen holder for rapid surgical and biopsy specimen imaging. Biomed Opt Express, BOE. 2024;15(5):2898–2909. doi:10.1364/BOE.522379

10. Ching-Roa VD, Huang CZ, Tang X, Heckman C, Ibrahim S, Giacomelli MG. Vacuum-based specimen holder for rapid surgical specimen imaging. Biomed Opt Express, BOE. 2025;16(6):2303–2311. doi:10.1364/BOE.561064

11. Gareau DS. Feasibility of digitally stained multimodal confocal mosaics to simulate histopathology. JBO. 2009;14(3):034050. doi:10.1117/1.3149853

12. Bennàssar A, Vilata A, Puig S, Malvehy J. Ex vivo fluorescence confocal microscopy for fast evaluation of tumour margins during Mohs surgery. British Journal of Dermatology. 2014;170(2):360–365. doi:10.1111/bjd.12671

13. Tri-modal confocal mosaics detect residual invasive squamous cell carcinoma in Mohs surgical excisions. Accessed August 14, 2025. https://www.spiedigitallibrary.org/journals/journal-of-biomedical-optics/volume-17/issue-06/066018/Tri-modal-confocal-mosaics-detect-residual-invasive-squamous-cell-carcinoma/10.1117/1.JBO.17.6.066018.full

14. Giacomelli MG, Husvogt L, Vardeh H, et al. Virtual Hematoxylin and Eosin Transillumination Microscopy Using Epi-Fluorescence Imaging. PLoS One. 2016;11(8):e0159337. doi:10.1371/journal.pone.0159337

15. Grizzetti L, Kuonen F. Ex vivo confocal microscopy for surgical margin assessment: A histology-compared study on 109 specimens. Skin Health and Disease. 2022;2(2):e91. doi:10.1002/ski2.91

16. Giacomelli MG, Faulkner-Jones BE, Cahill LC, Yoshitake T, Do D, Fujimoto JG. Comparison of nonlinear microscopy and frozen section histology for imaging of Mohs surgical margins. Biomed Opt Express, BOE. 2019;10(8):4249–4260. doi:10.1364/BOE.10.004249

17. Huang C, Ching-Roa V, Liu Y, Giacomelli MG. High-speed mosaic imaging using scanner-synchronized stage position sampling. JBO. 2022;27(1):016502. doi:10.1117/1.JBO.27.1.016502

18. Ching-Roa VD, Huang CZ, Ibrahim SF, Smoller BR, Giacomelli MG. Real-time Analysis of Skin Biopsy Specimens With 2-Photon Fluorescence Microscopy. JAMA Dermatology. 2022;158(10):1175–1182. doi:10.1001/jamadermatol.2022.3628

19. Huang CZ, Montague JE, Ching-Roa VD, Drage MG, Ibrahim SF, Giacomelli MG. Rapid clearing and imaging of Mohs and melanoma surgical margins using a low-cost tissue processor. Biomed Opt Express, BOE. 2024;15(2):700–714. doi:10.1364/BOE.510132

20. Giacomelli MG. Evaluation of silicon photomultipliers for multiphoton and laser scanning microscopy. JBO. 2019;24(10):106503. doi:10.1117/1.JBO.24.10.106503

21. Ching-Roa VD, Olson EM, Ibrahim SF, Torres R, Giacomelli MG. Ultrahigh-speed point scanning two-photon microscopy using high dynamic range silicon photomultipliers. Sci Rep. 2021;11(1):5248. doi:10.1038/s41598-021-84522-0

22. McHugh ML. Interrater reliability: the kappa statistic. Biochem Med (Zagreb). 2012;22(3):276–282.

23. Ashby D. Practical statistics for medical research. Douglas G. Altman, Chapman and Hall, London, 1991. No. of pages: 611. Price: £32.00. doi:10.1002/sim.4780101015

24. Marcelus H, Packert D. Addressing the histotechnologist shortage through improved classification and recognition. Journal of Histotechnology. 2024;47(4):143–145. doi:10.1080/01478885.2024.2424049

25. Davis MJ, Tegnander AN, McKay C, Nehal K. National Laboratory Technician Workforce Shortages: Implications for Mohs Micrographic Surgery. Dermatologic Surgery. 2024;50(9):876. doi:10.1097/DSS.0000000000004214

26. Heckman CM, Huang CZ, Ching-Roa VD, Ibrahim SF, Giacomelli MG. Two-photon microscopy allows for rapid imaging and diagnosis of Mohs specimens. JAAD Case Rep. 2025;67:63–67. doi:10.1016/j.jdcr.2025.10.042

